# High-risk human papillomavirus detection across the cervical disease spectrum in Abuja, Nigeria: a comparative cross-sectional study

**DOI:** 10.64898/2026.11.17.26368358

**Authors:** Juliet Oluchukwu Offor, Ernest Orji, George Uchenna Eleje, Imran O. Morhason-Bello, Ayodele Omotoso

## Abstract

Cervical cancer remains a major public health problem in Nigeria, but data on high-risk human papillomavirus (hrHPV) across the cervical disease spectrum are scarce for Abuja. We conducted a hospital-based comparative cross-sectional study of 101 women aged 25 years or older at Federal Medical Centre, Abuja: 59 with negative cytology (NILM) and 42 with cervical lesions (23 low- grade squamous intraepithelial lesions [LSIL], 8 high-grade lesions [HSIL], and 11 invasive cancers). Cervical specimens were tested using the Atila ScreenFire assay, which reports four pooled hrHPV target groups. hrHPV DNA was detected in 37/101 women (36.6%): 30.5% (18/59) in NILM, 26.1% (6/23) in LSIL, 50.0% (4/8) in HSIL, and 81.8% (9/11) in invasive cancer (Fisher–Freeman–Halton exact p = 0.005). Compared with NILM, invasive cancer had approximately tenfold higher odds of hrHPV detection (unadjusted odds ratio [OR] 10.25, 95% CI 2.01–52.28; age-adjusted OR 9.90, 95% CI 1.87–52.38). The HPV31/33/35/52/58 target group was detected most often (12.9%), followed by HPV16 and HPV18/45 (10.9% each) and HPV39/51/56/59/68 (5.9%). HPV16 was the most frequent target in invasive cancer (5/11). No target group was independently associated with lesion status. The NILM and lesion groups differed substantially in ethnicity, religion, and previous cervical screening, indicating different referral and screening pathways; the NILM group was not population-based. These findings show marked hrHPV enrichment in invasive cancer and substantial detection of non-16/18 target groups. They support HPV-based screening, genotype surveillance, and larger multicenter studies with population-based comparison groups.

## Introduction

Cervical cancer remains a major public health challenge in Nigeria. It is among the most common cancers affecting Nigerian women, with substantial mortality and persistent gaps in screening and timely treatment [1,2]. At Federal Medical Centre (FMC), Abuja, a 2018–2023 retrospective series found cervical cancer to be the most common gynecological malignancy, accounting for 39.66% of gynecological cancers; ovarian cancer contributed the largest share of deaths in that series [3]. Although HPV vaccination, cervical screening, and treatment of precancerous lesions are effective, cervical cancer continues to impose a substantial burden in Nigeria [1,2].

Globally, cervical cancer is the fourth most common cancer among women. In 2022, approximately 660,000 new cases and 350,000 deaths were estimated worldwide, with the majority of deaths occurring in low- and middle-income countries [4]. The World Health Organization (WHO) has established a global strategy for cervical cancer elimination based on high HPV vaccination coverage, effective screening of women at risk, and timely management of cervical precancer and invasive disease [4,5]. Achieving these targets requires epidemiological information that is relevant to individual populations and health systems.

Persistent infection with oncogenic HPV is the necessary cause of almost all cervical cancers [4,6]. More than 200 HPV types have been identified, of which approximately 40 infect the anogenital tract and a subset are classified as high-risk or carcinogenic because of their established association with cervical and other anogenital cancers [6,7]. Most HPV infections are transient and are cleared by the host immune response, whereas persistent infection with a carcinogenic HPV type may lead to the development of cervical precancer over 15–20 years [2,6]. The likelihood of persistence and progression is influenced by several host, behavioural, and environmental factors, including immunosuppression, HIV infection, cigarette smoking, reproductive factors, and other characteristics of the sexual and reproductive history [4,6].

HPV16 and HPV18 are responsible for a substantial proportion of cervical cancers globally and have therefore been central to the development of prophylactic HPV vaccines [8]. However, the distribution of carcinogenic HPV types varies between geographical regions and populations [9–12]. Nigerian studies have reported the circulation of several high-risk HPV types in addition to HPV16 and HPV18, including HPV31, HPV35, HPV45, HPV52, HPV56, and HPV58 [9–12]. Such geographical variation is relevant to cervical cancer prevention because it provides information about the HPV types circulating within populations and can inform interpretation of HPV-based screening and vaccination programmes.

Nigeria introduced HPV vaccination into the national immunisation programme in 2023, initially targeting girls aged 9–14 years [13]. The programme represents an important component of the country’s cervical cancer prevention strategy. Early implementation data have demonstrated substantial uptake among eligible girls in participating states, although coverage remains below the level required for population-wide elimination [14]. Because vaccination primarily targets HPV types responsible for a substantial proportion of cervical cancers, continued surveillance of circulating HPV types remains important for evaluating population-level prevention strategies. Screening coverage in Nigeria also remains limited and is predominantly opportunistic [1,2].

Studies conducted in different parts of Nigeria have reported considerable variation in HPV prevalence and genotype distribution. These differences may reflect geographical variation, differences in participant characteristics, cervical disease spectrum, HIV prevalence, specimen collection methods, and laboratory assays [9–12,15]. Furthermore, many Nigerian studies have evaluated HPV infection either among women attending screening services or among women with invasive cervical cancer, while relatively fewer studies have simultaneously examined women with normal cytology, precancerous cervical lesions, and invasive cervical cancer using a consistent molecular testing approach. Data from Abuja remain relatively limited despite the city’s large and heterogeneous population and its role as an important referral centre [11,16].

Accordingly, this study aimed to determine the detection frequency and distribution of hrHPV DNA target groups among women with normal cytology (NILM), precancerous cervical lesions (LSIL and HSIL), and invasive cervical cancer in Abuja, Nigeria, and to examine the association between hrHPV detection and cervical disease category. Demographic and screening characteristics are described where they are necessary to assess comparability of the hospital-based groups and to interpret potential referral-related confounding.

## Materials and methods

### Study design and setting

This comparative cross-sectional study was conducted at the Gynecologic Oncology and Radiation Oncology clinics and the cervical cancer screening clinic of Federal Medical Centre (FMC), Abuja, Nigeria, between 3 March 2025 and 28 February 2026. FMC Abuja is a tertiary referral facility serving a diverse urban and peri-urban population within the Federal Capital Territory and neighbouring states. HPV DNA testing was performed at the HPV Consortium, National Institute for Maternal, Newborn and Child Health, College of Medicine, University College Hospital, Orita- Mefa, Ibadan, Oyo State, Nigeria.

### Study population and eligibility criteria

The study population comprised women aged 25 years and older attending the study clinics during the study period. Participants were categorised into two principal groups according to cervical cytology and histopathological findings: women with normal cervical cytology, defined as negative for intraepithelial lesion or malignancy (NILM), and women with cervical lesions, including low-grade squamous intraepithelial lesion (LSIL), high-grade squamous intraepithelial lesion (HSIL), and invasive cervical cancer. All participants were recruited prospectively.

Women were eligible if they were aged 25 years and older, had an intact cervix, and provided written informed consent. Women were excluded if they had undergone total hysterectomy or trachelectomy, had incomplete clinical or laboratory data, or were unable to provide informed consent.

The primary outcomes were the detection frequency and distribution of high-risk human papillomavirus (hrHPV) DNA target groups and their association with the severity of cervical lesions.

### Sample size determination

The minimum sample size was estimated with the formula for comparing two independent proportions [17].

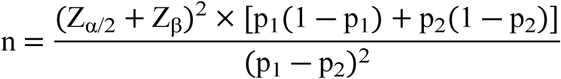

Using a two-sided 95% confidence level (Zα/2 = 1.96) and 90% power (Zβ = 1.282). The expected hrHPV prevalence was 3.5% in normal cytology (p1 = 0.035) and 28.8% in HSIL (p2 = 0.288), the HSIL prevalence being used as the expected prevalence for the combined lesion group [1]. The calculation gave 40 women per group (80 overall). A 5% allowance for non-evaluable participants raised this to 84, or 42 in each principal group, so the study was designed to compare NILM with the lesion group. In all, 101 women were enrolled. The minimum was exceeded partly because some women with an initially abnormal cytology result had NILM on histology. Of the 101, 59 had NILM and 42 had cervical lesions (23 LSIL, 8 HSIL, and 11 invasive cancer).

### Sampling technique

Consecutive sampling was employed. All eligible women presenting to the study clinics during the recruitment period were assessed for eligibility and enrolled prospectively. Recruitment into the individual lesion categories (LSIL, HSIL, and invasive cervical cancer) reflected the clinical case mix presenting to the study centre; therefore, predetermined equal sample sizes were not required for the individual lesion subgroups. Women with NILM were recruited from the same hospital clinics and therefore do not constitute a population-based comparison group.

### Data collection

Data were collected using a structured, interviewer-administered questionnaire adapted from previously validated instruments [18,19]. The questionnaire captured sociodemographic, reproductive, sexual, behavioural, and clinical characteristics, including age, marital status, educational attainment, ethnicity, religion, parity, age at sexual debut, lifetime number of sexual partners, oral contraceptive use, history of sexually transmitted infections, HIV status, previous cervical cancer screening, and knowledge of and previous vaccination against HPV. Ethnicity was recorded using the study questionnaire categories Hausa, Yoruba, Igbo, and Other, and religion as Christianity or Islam. These variables were used to describe the composition of the referral populations and potential confounding and were not interpreted as biological categories.

The questionnaire was pretested on about 10% of the calculated sample at a health facility with similar characteristics outside the study site, to assess clarity, comprehensibility, the sequence of questions, and completion time. Necessary modifications were made before the main study.

Cervical lesion status was determined from cytology (NILM) and/or histopathology (LSIL, HSIL, and invasive cancer), as clinically indicated; histopathology was the definitive basis for classifying premalignant and malignant lesions where it was available. The final classification was based on the diagnostic findings recorded during the study period and participants recruited prospectively into the strata group.

### Cervical specimen collection and HPV DNA testing

Trained clinicians collected exfoliated cervical cells with standard cervical brushes during routine pelvic examinations. Specimens were placed immediately into the transport medium supplied with the collection kit, transferred on dry ice to the FMC Abuja molecular laboratory, and stored at −40 to −80 °C until processing, in line with the manufacturer’s recommendations. Specimens that did not meet these storage and transport conditions were excluded.

High-risk HPV DNA testing was performed with the Atila ScreenFire assay, an isothermal amplification assay for 13 high-risk HPV genotypes that reports four target groups: HPV16; HPV18/45; HPV31/33/35/52/58; and HPV39/51/56/59/68. Commercially available HPV tests differ in the amount of genotype information they provide, and pooled-target assays such as ScreenFire report groups of types rather than individual genotypes [20]. A specimen was classified as having multiple-target positivity when more than one HPV target group was detected concurrently [21].

All specimen processing and laboratory procedures were performed in accordance with institutional standard operating procedures and quality-assurance protocols. Laboratory personnel followed the manufacturer’s instructions for specimen handling, testing, and interpretation of assay results.

An earlier testing round was not used for the analyses reported here because it did not yield results considered sufficiently reliable for study reporting. Following approval of an ethics amendment, the stored specimens were retested with the Atila ScreenFire assay at the HPV Consortium in Ibadan. Only ScreenFire results are reported in this manuscript.

To reduce recruitment-related selection bias within the participating clinics, consecutive sampling was used. All specimens contributing to the reported HPV analyses were assessed with the same assay and laboratory workflow. Because women with NILM and women with cervical lesions entered the study through different screening and referral pathways, between-group demographic and screening differences were examined descriptively and are explicitly considered when interpreting the comparisons.

### Statistical analysis

Data were coded, entered, cleaned, and analysed using IBM SPSS Statistics version 31.0 (IBM Corp., Armonk, NY, USA) [22]. Continuous variables were assessed for distribution and summarised as means with standard deviations (SD) for approximately normally distributed data or medians with interquartile ranges (IQR) for non-normally distributed data. Categorical variables were summarised using frequencies and percentages, with 95% confidence intervals for proportions calculated by the Wilson method.

Associations between categorical variables, including the overall association between lesion category and hrHPV detection, were evaluated using the Pearson chi-square test or Fisher’s exact test, as appropriate. The overall association between lesion category and hrHPV detection was assessed with the Fisher–Freeman–Halton exact test because several expected cell counts were below 5. Continuous variables were compared using the Mann–Whitney U test. Univariable logistic regression analyses were initially performed to assess associations between potential explanatory variables and HPV positivity or cervical lesion status. Variables meeting the prespecified criteria for inclusion were subsequently considered in multivariable logistic regression models. Because only 11 women had invasive cancer and 42 had any lesion, multivariable models are exploratory, and their estimates are imprecise. Associations were reported as odds ratios (ORs) with 95% confidence intervals (CIs); unadjusted ORs and CIs were calculated from 2 × 2 tables by the Woolf (logit) method, and adjusted ORs by logistic regression. All statistical tests were two-sided, and a p-value ≤0.05 was considered statistically significant. Where multiple exploratory comparisons were conducted, the Benjamini– Hochberg procedure was applied to control the false discovery rate.

Analyses were conducted using available-case data for variables with missing observations; no imputation was performed.

### Ethics statement

Ethical approval was obtained from the Health Research Ethics Committee of Federal Medical Centre, Abuja, before the study began (protocol number FMCABJ/HREC/2025/223; approval period 1 March 2025 to 28 February 2026; National Health Research Ethics Committee registration number NHREC/FMC-JABI-HREC/03/03/2023B). A later amendment, approved by the same committee, covered retesting of the stored specimens at the HPV Consortium in Ibadan; it involved no additional specimens or procedures. Written informed consent was obtained from all participants before enrolment. Participation was voluntary, and participants were informed of their right to withdraw from the study at any time without affecting their clinical care.

Participant confidentiality was maintained throughout the study. Unique study identification numbers were used in place of personal identifiers, and identifying information was kept separate from laboratory and analytical datasets. The study was conducted in accordance with the principles of the Declaration of Helsinki and applicable Nigerian national ethical and regulatory requirements.

### Reporting guideline

This study was reported in accordance with the Strengthening the Reporting of Observational Studies in Epidemiology (STROBE) guidelines for cross-sectional studies [23] (S1 Checklist).

## Results

### Participant flow

Of 118 women assessed for eligibility, 17 were excluded (11 did not meet the eligibility criteria, 4 declined or did not give consent, and 2 had undergone total hysterectomy). The remaining 101 women were enrolled and tested with the Atila ScreenFire assay: 59 with NILM and 42 with cervical lesions (23 LSIL, 8 HSIL, and 11 invasive cancer) (Fig 1).

**Fig 1.**
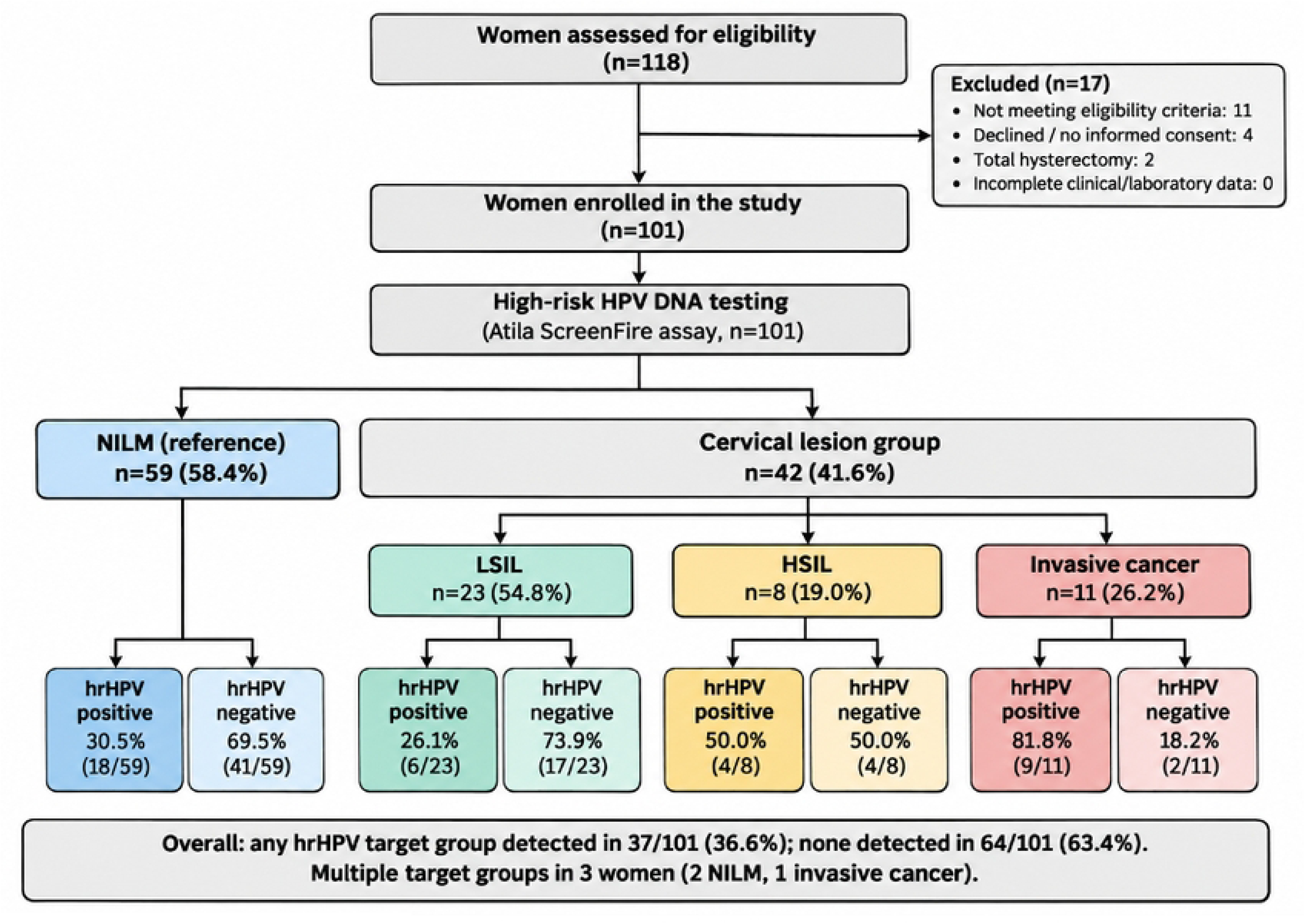
Participant flow through the study. The diagram shows women assessed, exclusions, enrolment, cervical disease categories, and hrHPV detection within each category. Percentages in the terminal boxes are the proportions with or without at least one hrHPV target detected. NILM, negative for intraepithelial lesion or malignancy; LSIL, low-grade squamous intraepithelial lesion; HSIL, high-grade squamous intraepithelial lesion; hrHPV, high-risk human papillomavirus.

### Participant characteristics

A total of 101 women were included: 59 with NILM and 42 with cervical lesions (23 LSIL, 8 HSIL, and 11 invasive cervical cancer). The overall mean age was 46.9 ± 12.9 years. Baseline sociodemographic and reproductive characteristics are summarised in Table 1. The NILM and lesion groups differed markedly in ethnicity and religion (Hausa: 67.8% vs 4.8%; Muslim: 71.2% vs 19.0%; both p < 0.0001). Previous cervical screening was also far more common in the lesion group (71.4%, 30/42) than in the NILM group (23.7%, 14/59; Fisher’s exact p < 0.0001). These contrasts probably reflect the different referral and screening pathways that feed the study clinics, and they mean that the NILM group should not be read as a population-based comparison.

**Table 1.** Baseline sociodemographic and reproductive characteristics of participants by cervical disease category.

| **Characteristic** | **Total (N=101)** | **NILM (n=59)** | **LSIL (n=23)** | **HSIL (n=8)** | **Cancer (n=11)** |
|---|---|---|---|---|---|
| Age, years, mean ± SD | 46.9 ± 12.9 | 44.6 ± 12.6 | 48.6 ± 13.4 | 49.0 ± 8.8 | 53.5 ± 14.1 |
| Age, years, median (IQR) | 44 (37–55) | 41 (35–55) | 45 (39–54) | 51 (47–53) | 51 (44–60) |
| Age group, n (%) |  |  |  |  |  |
| ≤30 | 9 (8.9) | 8 (13.6) | 1 (4.3) | 0 (0.0) | 0 (0.0) |
| 31–40 | 29 (28.7) | 20 (33.9) | 6 (26.1) | 2 (25.0) | 1 (9.1) |
| 41–50 | 27 (26.7) | 13 (22.0) | 8 (34.8) | 2 (25.0) | 4 (36.4) |
| 51–60 | 20 (19.8) | 9 (15.3) | 5 (21.7) | 3 (37.5) | 3 (27.3) |
| >60 | 16 (15.8) | 9 (15.3) | 3 (13.0) | 1 (12.5) | 3 (27.3) |
| Marital status, n (%) |  |  |  |  |  |
| Married | 70 (69.3) | 39 (66.1) | 17 (73.9) | 6 (75.0) | 8 (72.7) |
| Widowed | 14 (13.9) | 9 (15.3) | 2 (8.7) | 1 (12.5) | 2 (18.2) |
| Divorced | 9 (8.9) | 7 (11.9) | 1 (4.3) | 1 (12.5) | 0 (0.0) |
| Single | 8 (7.9) | 4 (6.8) | 3 (13.0) | 0 (0.0) | 1 (9.1) |
| Education, n (%) |  |  |  |  |  |
| None | 11 (10.9) | 11 (18.6) | 0 (0.0) | 0 (0.0) | 0 (0.0) |
| Primary | 9 (8.9) | 6 (10.2) | 0 (0.0) | 1 (12.5) | 2 (18.2) |
| Secondary | 26 (25.7) | 19 (32.2) | 2 (8.7) | 1 (12.5) | 4 (36.4) |
| Tertiary | 52 (51.5) | 23 (39.0) | 20 (87.0) | 6 (75.0) | 3 (27.3) |
| Missing | 3 (3.0) | 0 (0.0) | 1 (4.3) | 0 (0.0) | 2 (18.2) |
| Ethnicity, n (%) |  |  |  |  |  |
| Hausa | 42 (41.6) | 40 (67.8) | 0 (0.0) | 1 (12.5) | 1 (9.1) |
| Igbo | 29 (28.7) | 11 (18.6) | 13 (56.5) | 2 (25.0) | 3 (27.3) |
| Yoruba | 7 (6.9) | 1 (1.7) | 3 (13.0) | 3 (37.5) | 0 (0.0) |
| Other | 23 (22.8) | 7 (11.9) | 7 (30.4) | 2 (25.0) | 7 (63.6) |
| Occupation, n (%) |  |  |  |  |  |
| Self-employed | 45 (44.6) | 29 (49.2) | 7 (30.4) | 5 (62.5) | 4 (36.4) |
| Civil servant | 33 (32.7) | 15 (25.4) | 12 (52.2) | 3 (37.5) | 3 (27.3) |
| Housewife | 15 (14.9) | 11 (18.6) | 0 (0.0) | 0 (0.0) | 4 (36.4) |
| Retired | 5 (5.0) | 1 (1.7) | 4 (17.4) | 0 (0.0) | 0 (0.0) |
| Unemployed/Other | 3 (3.0) | 3 (5.1) | 0 (0.0) | 0 (0.0) | 0 (0.0) |
| Religion, n (%) |  |  |  |  |  |
| Christianity | 51 (50.5) | 17 (28.8) | 23 (100.0) | 4 (50.0) | 7 (63.6) |
| Islam | 50 (49.5) | 42 (71.2) | 0 (0.0) | 4 (50.0) | 4 (36.4) |
| Income level, n (%) |  |  |  |  |  |
| Low | 45 (44.6) | 31 (52.5) | 5 (21.7) | 2 (25.0) | 7 (63.6) |
| Medium | 52 (51.5) | 26 (44.1) | 18 (78.3) | 6 (75.0) | 2 (18.2) |
| High | 4 (4.0) | 2 (3.4) | 0 (0.0) | 0 (0.0) | 2 (18.2) |
| Age at first sexual intercourse, years, mean ± SD | 20.1 ± 3.4 | 19.9 ± 3.5 | 21.1 ± 3.6 | 19.2 ± 4.1 | 19.5 ± 2.3 |
| Lifetime sexual partners, median (IQR) | 1 (1–3) | 1 (1–3) | 2 (1–4) | 2 (1–3) | 1 (1–2) |
| Parity, median (IQR) | 4 (2–6) | 4 (2–6) | 3 (2–3) | 3 (2–4) | 4 (4–6) |
| History of STI, n (%) |  |  |  |  |  |
| Yes | 29 (28.7) | 15 (25.4) | 6 (26.1) | 2 (25.0) | 6 (54.5) |
| No | 72 (71.3) | 44 (74.6) | 17 (73.9) | 6 (75.0) | 5 (45.5) |
| Previous cervical cancer screening, n (%) |  |  |  |  |  |
| Yes | 44 (43.6) | 14 (23.7) | 19 (82.6) | 5 (62.5) | 6 (54.5) |
| No | 57 (56.4) | 45 (76.3) | 4 (17.4) | 3 (37.5) | 5 (45.5) |

\*Notes: Percentages are column percentages within each cervical disease category. Comparisons of NILM with the lesion group mentioned in the text used the Pearson χ² or Fisher’s exact test, as appropriate. Other ethnicities: Edo, Ebira, Tiv, Fulani, Urhobo, Igala, Bette, Eggon, Igede, Idoma, Kabba, Kanuri, Gbaji, and Bungudu.\*

### Distribution of high-risk HPV assay target groups

Overall target-group frequencies, which include women with more than one target group, were HPV31/33/35/52/58 (13/101; 12.9%), HPV16 (11/101; 10.9%), HPV18/45 (11/101; 10.9%), and HPV39/51/56/59/68 (6/101; 5.9%). Three women had multiple-target positivity: two in the NILM group and one with invasive cancer. Exact mutually exclusive assay positivity patterns are shown in S1 Fig.

### High-risk HPV DNA detection by cervical lesion category

Detection rates across the disease spectrum were 30.5% (18/59) in NILM, 26.1% (6/23) in LSIL, 50.0% (4/8) in HSIL, and 81.8% (9/11) in invasive cervical cancer (Fig 2). The overall association between lesion category and hrHPV positivity was statistically significant (Fisher–Freeman– Halton exact p = 0.005; Pearson χ² p = 0.006, which is less reliable here because several expected counts were below 5). The pattern was driven primarily by invasive cancer: positivity among women with any lesion (45.2%, 19/42) did not differ significantly from NILM (Fisher’s exact p = 0.147), and pairwise comparisons of LSIL or HSIL with NILM were not significant (p = 0.79 and p = 0.42, respectively). The pattern was not monotonic, because positivity was slightly lower in LSIL than in NILM.

**Fig 2.**
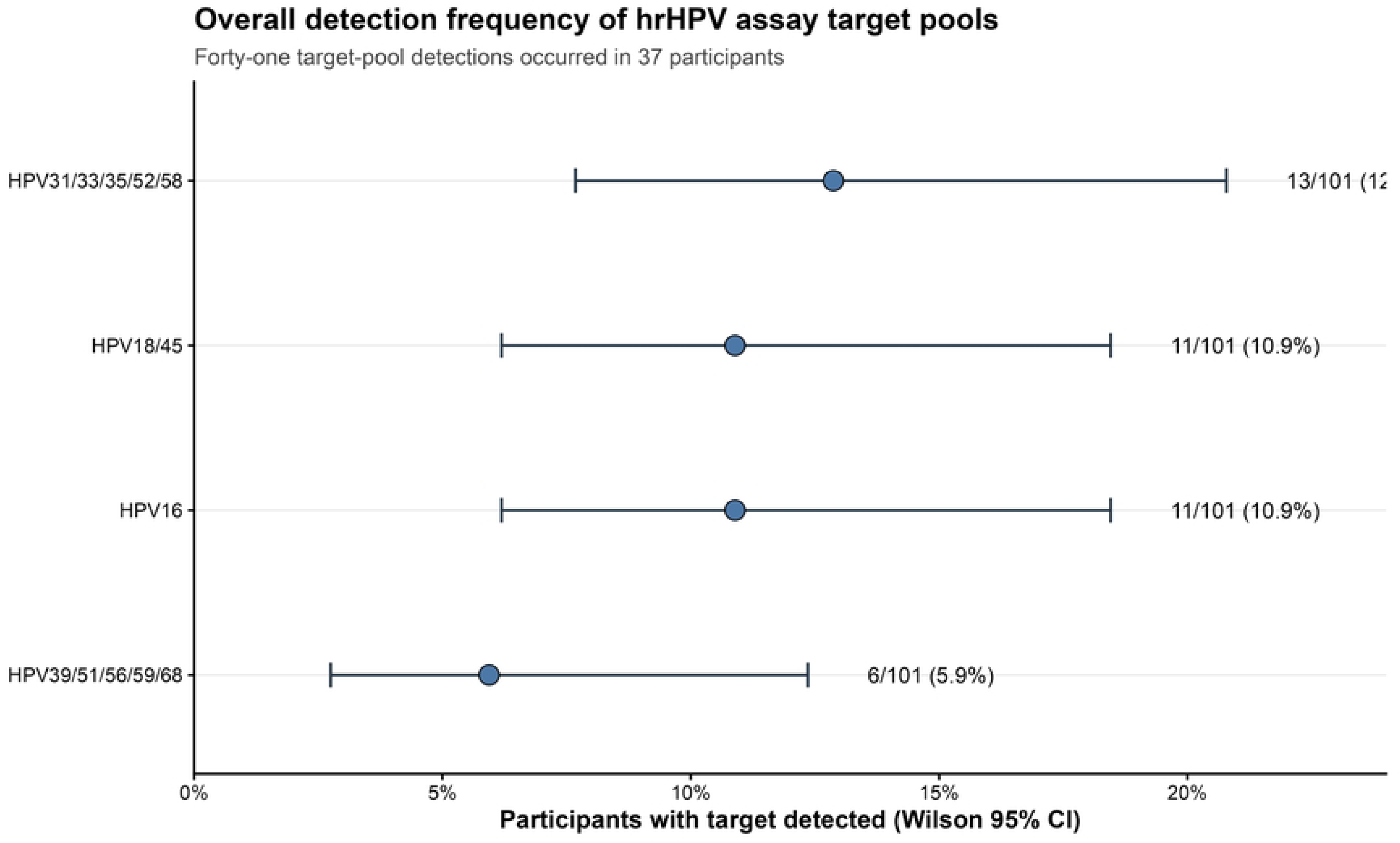
High-risk HPV detection by cervical disease category. Bars show the percentage of participants with at least one hrHPV target detected, with Wilson 95% confidence intervals; labels show positive/total and percentage. Detection was highest in invasive cervical cancer. Category-specific hrHPV detection estimates, including Wilson 95% confidence intervals, are shown in Table 2.

**Table 2.** High-risk HPV DNA detection by cervical lesion category.

| **Group** | **n** | **Age, mean (SD)** | **HPV positive** | **Percent** | **Wilson 95% CI** | **Multiple targets** |
|---|---|---|---|---|---|---|
| NILM | 59 | 44.6 (12.6) | 18/59 | 30.5% | 20.3–43.1% | 2 |
| LSIL | 23 | 48.6 (13.4) | 6/23 | 26.1% | 12.5–46.5% | 0 |
| HSIL | 8 | 49.0 (8.8) | 4/8 | 50.0% | 21.5–78.5% | 0 |
| Invasive cancer | 11 | 53.5 (14.1) | 9/11 | 81.8% | 52.3–94.9% | 1 |
| Overall | 101 | 46.9 (12.9) | 37/101 | 36.6% | 27.9–46.4% | 3 |

Compared with NILM, invasive cervical cancer was strongly associated with hrHPV detection (unadjusted OR 10.25, 95% CI 2.01–52.28; Fisher’s exact p = 0.002). In an age-adjusted model restricted to women with invasive cancer and NILM (n = 70), the association remained significant (adjusted OR 9.90, 95% CI 1.87–52.38; p = 0.007). Age itself was not independently associated with malignancy in this model (adjusted OR per 10 years 1.63, 95% CI 0.93–2.85; p = 0.086), but the model contained only 11 cancers and was underpowered for this comparison. The unadjusted disease-category odds ratios are shown in Fig 3.

**Fig 3.**
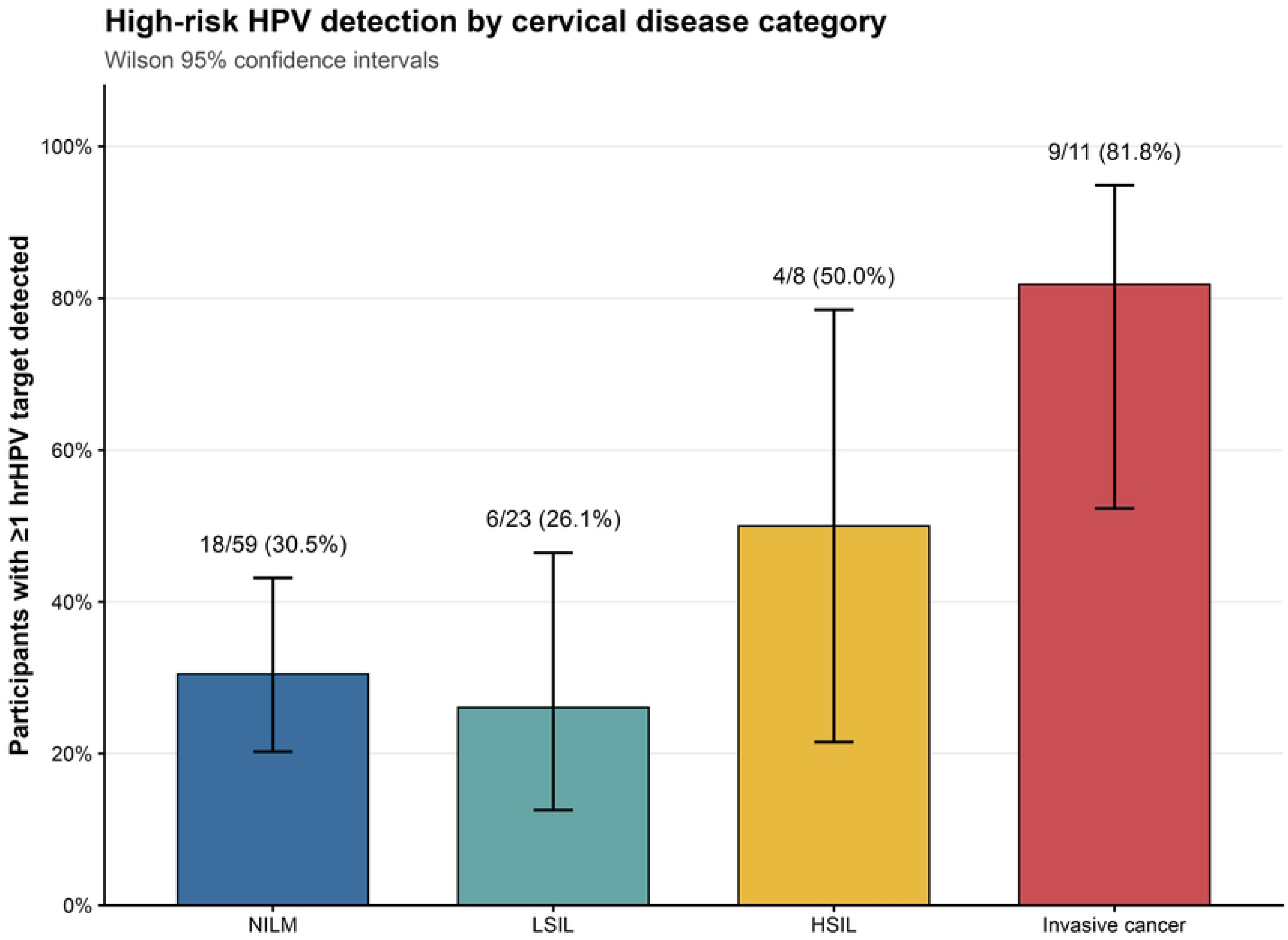
Association between hrHPV detection and cervical disease category. Forest plot of unadjusted odds ratios with 95% confidence intervals for LSIL, HSIL, and invasive cervical cancer compared with NILM. The dashed vertical line marks an odds ratio of 1; the horizontal axis is logarithmic. *Note: Multiple-target positivity was uncommon, occurring in only three women (two with NILM and one with invasive cancer). Wilson 95% confidence intervals are shown for proportions.

### Distribution of individual target groups across lesion categories

HPV16 was the most frequently detected target among women with invasive cancer (5/11, 45.5%). In contrast, the HPV31/33/35/52/58 pool was the most common target among women with NILM and LSIL. Fig 4 presents target-pool detection across cervical disease categories; participants with multiple positive pools contribute to each applicable target-pool row. Overall target-pool detection frequencies with Wilson 95% confidence intervals are summarized in Fig 5.

**Fig 4.**
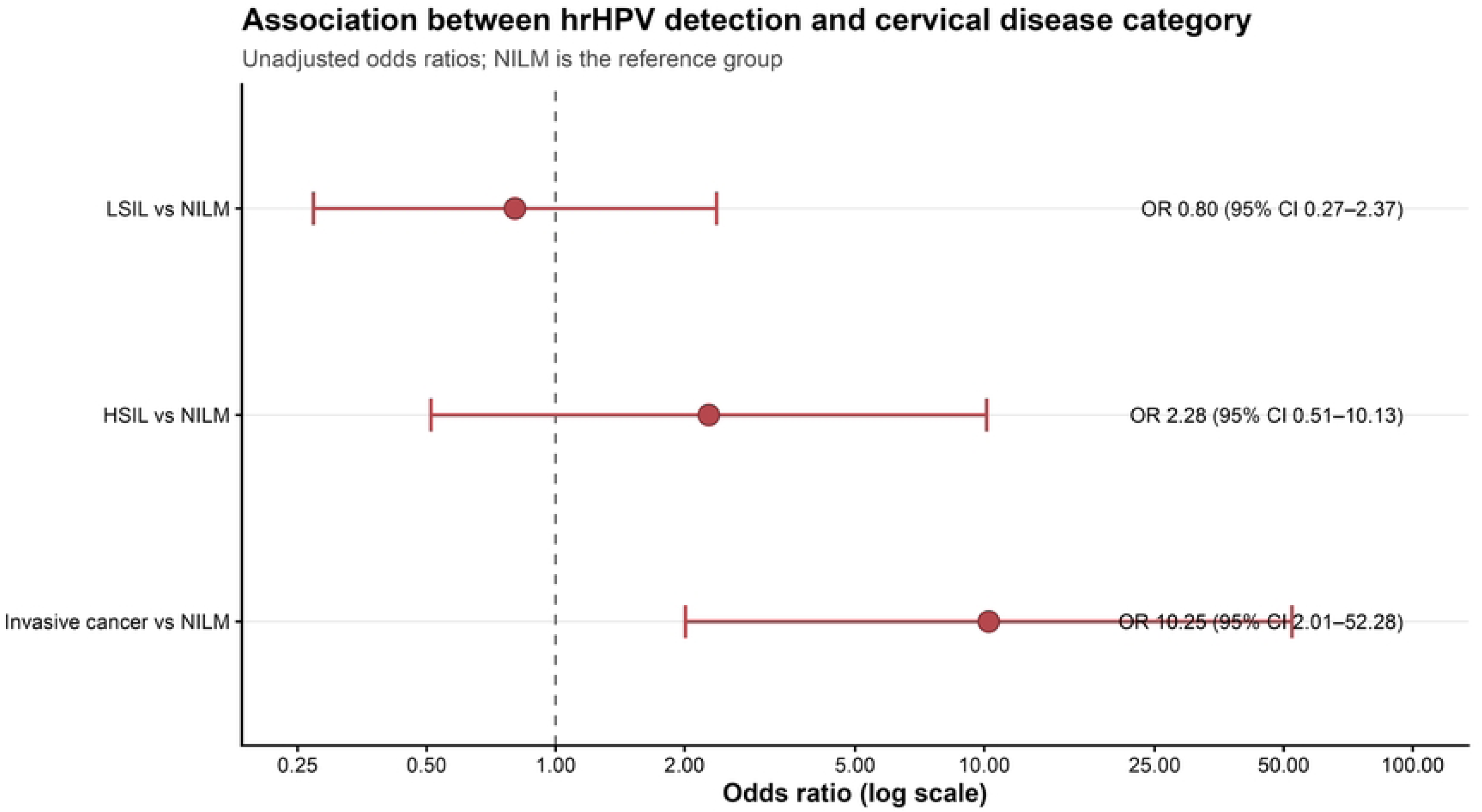
hrHPV target-pool detection by cervical disease category. Heatmap cells show positive/total and percentage for each ScreenFire target pool within NILM, LSIL, HSIL, and invasive cervical cancer. Participants with multiple positive target pools contribute to each applicable row.

**Fig 5.**
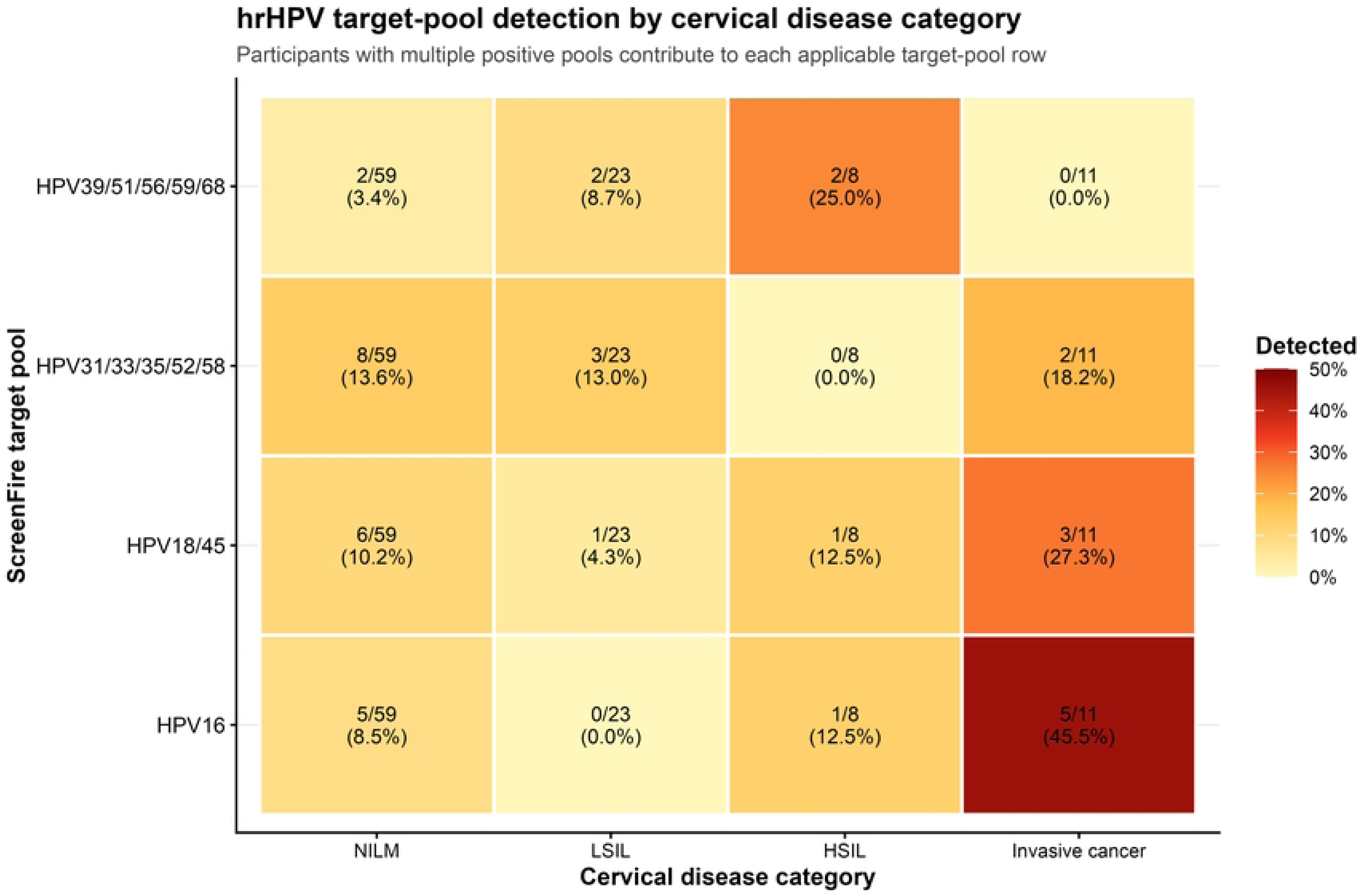
Overall detection frequency of hrHPV assay target pools. Points show the percentage of all 101 participants with each target pool detected; horizontal bars are Wilson 95% confidence intervals. Target-pool detections are not mutually exclusive: 41 target-pool detections occurred in 37 participants.

### Target groups and the presence of cervical lesions

To assess whether any target group was associated with lesion status independently of other factors, we fitted an exploratory multivariable logistic regression model for any lesion (LSIL, HSIL, or invasive cancer) versus NILM. The four target groups were entered together with age, HIV status, number of children, oral contraceptive use, and age at first sexual intercourse. Complete-case analysis included 99 women (41 with a lesion and 58 with NILM) (Table 3; full model output in S2 Table).

**Table 3.** Association of the four hrHPV target groups with the presence of a cervical lesion (LSIL, HSIL, or invasive cancer) versus NILM in an exploratory complete-case multivariable model (n = 99)

| **Variable** | **OR** | **95% CI lower** | **95% CI upper** | **p-value** |
|---|---|---|---|---|
| HPV16 | 2.270 | 0.529 | 9.732 | 0.270 |
| HPV18/45 | 1.376 | 0.335 | 5.661 | 0.658 |
| HPV31/33/35/52/58 | 0.704 | 0.170 | 2.918 | 0.629 |
| HPV39/51/56/59/68 | 5.252 | 0.767 | 35.957 | 0.091 |

\*Notes: OR = odds ratio; CI = confidence interval. Odds ratios are adjusted for the other target groups, age, HIV status, number of children, oral contraceptive use, and age at first sexual intercourse; full estimates for all covariates are reported in S2 Table. The model was not adjusted for ethnicity or religion despite marked between-group imbalance, and estimates are imprecise because of the limited number of events.\*

The inability of the assay to resolve individual genotypes within each pool, together with the small number of malignant cases (n = 11), limited the power to detect genotype-specific associations. Overall hrHPV positivity was associated with invasive cancer, whereas no single target group was independently associated with lesion status.

### Exploratory associations with any HPV positivity

Univariable screening of demographic, behavioural, and clinical variables against any HPV positivity identified a nominal association with history of sexually transmitted infection (p = 0.022). After Benjamini–Hochberg adjustment for multiple testing, this association was no longer significant (adjusted p = 0.348). No other variable examined remained significant after correction (Table 4).

**Table 4.** Exploratory associations with any HPV positivity.

| **Variable** | **Complete n** | **Test p** | **BH-adjusted p** |
|---|---|---|---|
| STI history | 101 | 0.022 | 0.348 |
| Age at first sex | 99 | 0.124 | 0.542 |
| Number of children | 101 | 0.083 | 0.542 |
| Age at first pregnancy | 93 | 0.135 | 0.542 |
| Income level | 101 | 0.171 | 0.549 |
| Knows HPV is cervical cancer risk | 101 | 0.217 | 0.579 |
| HIV documented | 101 | 0.256 | 0.586 |
| Marital status | 101 | 0.325 | 0.651 |
| Previous cancer screening | 101 | 0.533 | 0.856 |
| Age | 101 | 0.535 | 0.856 |
| HPV vaccination | 101 | 0.622 | 0.905 |
| Education | 98 | 0.734 | 0.956 |

\*Note: These tests screen demographic, behavioural, and clinical variables one at a time. They are hypothesis-generating; sparse categories, self-report, missingness, and multiple testing limit interpretation. Benjamini–Hochberg (BH) adjustment was applied across 16 variables screened; four variables with unadjusted p ≥ 0.73 are not shown.\*

## Discussion

This comparative cross-sectional study assessed hrHPV DNA target groups among women with normal cytology, precancerous cervical lesions, and invasive cervical cancer at a tertiary centre in Abuja, Nigeria. Among 101 women, 37 (36.6%) had at least one hrHPV target detected. Detection was 30.5% in NILM, 26.1% in LSIL, 50.0% in HSIL, and 81.8% in invasive cancer. The pattern was not strictly monotonic because LSIL positivity was slightly lower than NILM positivity, but detection was substantially higher in HSIL and, particularly, in invasive cancer. Women with invasive cancer had approximately tenfold higher odds of hrHPV detection than women with NILM (OR 10.25, 95% CI 2.01–52.28; p = 0.002), with a similar estimate after age adjustment (adjusted OR 9.90, 95% CI 1.87–52.38; p = 0.007). The wide intervals reflect the small number of cancers (n = 11).

The overall hrHPV detection of 36.6% reflects the case mix of this hospital-based sample (42% with cervical lesions) and is not a population prevalence estimate; it lies within the range of previous Nigerian estimates, although those studies recruited different populations [11,12]. A systematic review and meta-analysis reported a pooled HPV prevalence of approximately 32% among Nigerian women [15], while another systematic review estimated pooled cervical HPV prevalence at 20.65%, with substantial geographical heterogeneity [9]. The present estimate is also similar to the 37% reported among women in Abuja by Akarolo-Anthony et al. [11], but considerably higher than the 11.8% reported among apparently healthy women in Kaduna [25]. Differences between studies may reflect variation in participant characteristics, clinical setting, geographical location, genotype panels, and laboratory methods. The 30.5% positivity among women with normal cytology is also high relative to the 3.5% assumed in the sample size calculation and probably reflects, at least in part, the hospital-based recruitment of the NILM group [1].

The most notable finding was the high HPV detection among women with invasive cervical cancer (81.8%). This is consistent with recent West African evidence, including a 2025 systematic review and meta-analysis reporting a pooled HPV prevalence of 81.4% among women with cervical cancer [26]. Similarly high prevalence has been reported in Ibadan, Nigeria [27]. Two of the 11 invasive cancers were HPV-negative with this assay. Possible explanations include HPV- independent cancers, loss of target sequences, or false-negative results with a pooled-target assay, but histological type was not analysed here, and no explanation can be confirmed. HPV detection was 50.0% among women with HSIL, compared with 26.1% in LSIL and 30.5% in NILM. The similar proportions in NILM and LSIL should be interpreted cautiously because HPV infection can occur in women with normal cytology and LSIL may represent a transient infection [6,24]; the LSIL subgroup was also small. The cross-sectional design does not permit inference regarding HPV persistence or progression from LSIL through HSIL to invasive cancer.

The HPV31/33/35/52/58 target group was the most frequently detected (12.9%), followed by HPV16 and HPV18/45 (10.9% each) and HPV39/51/56/59/68 (5.9%). This is consistent with Nigerian studies showing substantial circulation of non-16/18 oncogenic types [9,12]. Akarolo- Anthony et al. identified HPV35, HPV56, HPV58, HPV45, and HPV59 among the predominant types in Abuja [11], and studies from Lagos and southwestern Nigeria have reported substantial representation of HPV31, HPV35, and HPV16 [28,29]. The distribution of types in cervical lesions also varies across South Africa and sub-Saharan Africa [30,31]. Because the assay reports pooled groups, our data cannot indicate which individual types, such as HPV35 or HPV58, accounted for positivity in the HPV31/33/35/52/58 pool. This matters for vaccine planning: the nonavalent vaccine covers HPV31, 33, 45, 52, and 58 but not HPV35 [32], which was among the predominant types in an earlier Abuja study [11]; therefore, resolving this pool is a priority.

HPV16 was detected in 10.9% of participants, consistent with its well-established importance in cervical cancer [7,8]. However, HPV16 was not independently associated with the presence of cervical lesions in the multivariable model (OR 2.27, 95% CI 0.53–9.73; p = 0.270), and none of the target groups showed an independent association; the study was not powered to examine this. The HPV18/45 target was detected at the same overall frequency as HPV16, which is noteworthy because HPV45 has been reported to contribute substantially to cervical cancer in African populations [1]; the pooled assay, however, cannot distinguish HPV18 from HPV45. Multiple- target positivity was uncommon (three women), although comparison with other studies is limited by differences in assay design. The quadrivalent vaccine used when Nigeria introduced its national programme targets HPV16 and HPV18 [13], whereas several of the detections here fall in pools containing other types. These findings cannot assess vaccine effectiveness, but they support continued type surveillance and consideration of broader-spectrum vaccines such as the nonavalent vaccine.

The factors associated with lesion status other than HPV are analysed in the companion paper on risk factors. In brief, older age was independently associated with the presence of a lesion, oral contraceptive use showed an inverse association that should not be read as protective, and HIV status, parity, and age at first sexual intercourse were not independently associated; all of these estimates are imprecise and may be confounded by the ethnic and religious imbalance between the groups. We therefore interpret the present findings at the level of HPV detection and target groups and do not repeat the risk-factor analysis here.

The NILM and lesion groups differed in more than age. Hausa and Muslim women made up most of the NILM group (67.8% and 71.2%) but few of the lesion group (4.8% and 19.0%); tertiary education was reported by 74.4% of women with lesions (29/39 with data) and 39.0% of NILM women (23/59); and 71.4% of women with lesions, against 23.7% of those with NILM, had been screened before. These contrasts are unlikely to be biological. They more probably reflect how women reached the hospital: women with lesions included many who were already in the screening and referral system and were referred to the oncology clinics from across the Federal Capital Territory and neighbouring states, whereas the NILM group came largely from women attending for first-time or routine evaluation. Two points follow. First, a comparison of HPV positivity between these groups is partly a comparison between referral populations, which is another reason to read the 30.5% positivity in NILM as a hospital-based figure. Second, future studies in Abuja should sample across ethnic and religious groups, or adjust for them, and should record how women were referred. Our data cannot show whether prior screening differed by ethnicity or religion within groups. Awareness of HPV and vaccination among these women are reported separately.

Strengths of this study include the simultaneous evaluation of women with NILM, LSIL, HSIL, and invasive cancer in one clinical setting using a single molecular assay for all analysed results, consecutive recruitment, and detailed demographic, behavioural, and clinical data. Limitations include the modest sample size, particularly the small HSIL (n = 8) and invasive cancer (n = 11) groups, which produced wide confidence intervals and limited power for genotype-specific inference; a sample size calculation that assumed a much lower HPV prevalence among NILM women than was observed and was not designed for LSIL or cancer comparisons; the pooled nature of the ScreenFire targets, which precludes identification of individual genotypes; the cross-sectional design, which cannot assess persistence, clearance, or progression; hospital-based, single-centre recruitment, with a NILM group that is not population-based and that differed markedly from the cervical lesion group in ethnicity and religion, which could not be adjusted for in the exploratory models; lesion classification based on cytology and/or histopathology as clinically indicated, which may differ in accuracy across categories; self-reported behavioural variables subject to recall and social-desirability bias; storage of the specimens and an earlier unsuccessful round of testing before ScreenFire testing, although ScreenFire returned valid results for all 101 women; and exploratory multivariable models with few events.

Detection of hrHPV in more than one-third of participants and in more than four-fifths of women with invasive cancer is consistent with the WHO recommendation to use HPV DNA testing as the preferred primary screening method [5,33]. This study, however, did not evaluate screening test performance, and sensitivity and specificity cannot be inferred. The high positivity among women with normal cytology (30.5%) also illustrates that HPV positivity alone does not indicate disease and that triage strategies are needed [33]. The presence of several high-risk target groups supports continued genotype-resolving surveillance beyond HPV16 and HPV18. Nigerian cancer data using next-generation sequencing have similarly identified HPV16, 18, 45, 35, and 52 among the most prevalent high-risk types in cervical cancer [12]. These observations do not imply failure of the national vaccination programme, which this study was not designed to evaluate; rather, they highlight the value of continued genotype surveillance and the potential public-health advantage of broad vaccine coverage [8,13]. Integrated education, vaccination, and screening, delivered as complementary components of elimination strategies, remain essential [5,34,35].

### Conclusion

High-risk HPV DNA was detected across the cervical disease spectrum in this hospital-based sample from Abuja and was strongly enriched in invasive cancer (81.8%; approximately tenfold higher odds than in NILM). Target groups containing neither HPV16 nor HPV18/45 accounted for nearly half of all detections, and HPV16 led only in invasive cancer; supporting continued genotype-resolving surveillance and the potential value of broad HPV vaccine type coverage. Women with lesions and women with normal cytology also differed sharply in ethnicity, religion, and prior screening, indicating that the groups came through different referral and screening pathways; HPV positivity across them should be read as hospital-based and not as a population estimate. Three steps would turn these preliminary findings into evidence that can guide practice: genotype-resolving analysis of the HPV31/33/35/52/58 and HPV18/45 pools to establish which individual types, notably HPV35 and HPV45, account for the detections; larger multicenter studies with population-based comparison groups that sample or adjust for ethnicity, religion, and screening history; and follow-up of HPV-positive women with normal cytology to define persistence and progression. Until then, the results support HPV-based screening with triage and continued genotype surveillance in Nigeria.

## Data Availability

Participant-level clinical, behavioural, cervical-screening, and HPV data cannot be deposited publicly because the informed-consent terms and ethical approval for this small, single-centre clinical cohort restrict unrestricted public release of potentially sensitive participant data. De-identified data underlying the reported findings are available to qualified researchers upon reasonable request and approval by the Health Research Ethics Committee, Federal Medical Centre, Abuja. Requests for data access may be directed to. The authors do not make decisions regarding data access.

## Acknowledgements

The authors thank the women who participated in the study and the consultants, residents, laboratory personnel, and support staff of the Department of Obstetrics and Gynecology, Federal Medical Centre Abuja, for their contributions to participant recruitment, clinical care, and study implementation. Language-editing tools were used to improve grammar and presentation; the authors remain responsible for the scientific content.

## Supporting information captions

**S1 Fig..**
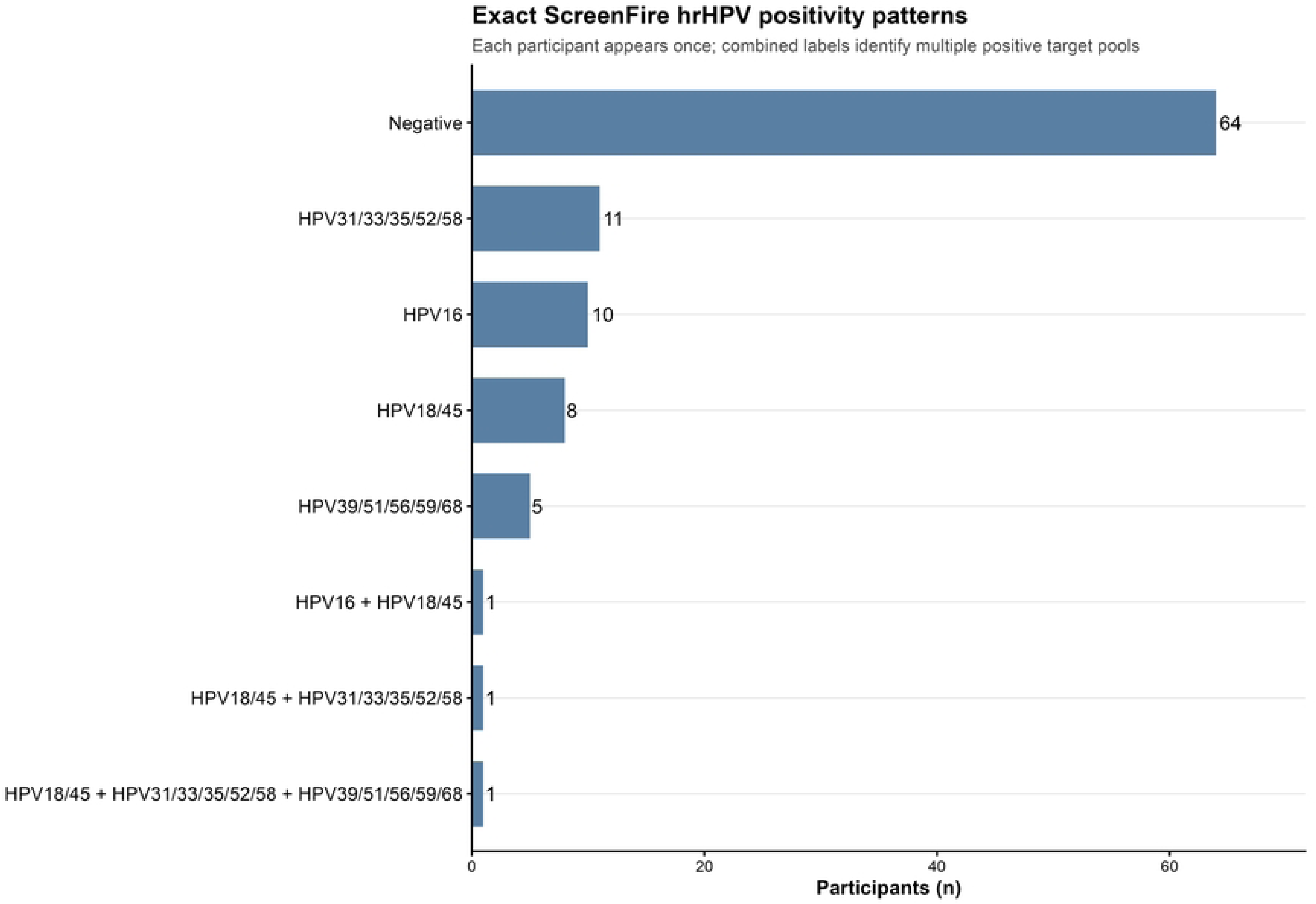
Exact ScreenFire hrHPV positivity patterns. Horizontal bars show mutually exclusive assay-result patterns among all 101 participants. Each participant appears once; combined labels identify participants with more than one positive target pool.

S1 Checklist. STROBE Statement—checklist of items that should be included in reports of cross-sectional studies.

S2 Table. Full logistic regression output for analyses reported in the manuscript. Model 1 is the age-adjusted association between any hrHPV detection and invasive cancer versus NILM (n = 70). Model 2 is the exploratory complete-case model for any cervical lesion versus NILM (n = 99), including all prespecified covariates and model-fit statistics.

## Notes

### Competing Interest Statement

The authors have declared no competing interest.

### Clinical Trial

NA

### Author Declarations

Ethical approval for this study was obtained from the Health Research Ethics Committee of the Federal Medical Centre, Abuja, Nigeria (Approval No. FMCABJ/HREC/2025/223). Written informed consent was obtained from all participants before enrolment in the study.

